# 6-Month Follow Up of 8679 Hospitalized COVID-19 Patients in Germany: A Nationwide Cohort Study

**DOI:** 10.1101/2021.04.24.21256029

**Authors:** Christian Günster, Reinhard Busse, Melissa Spoden, Tanja Rombey, Gerhard Schillinger, Wolfgang Hoffmann, Steffen Weber-Carstens, Andreas Schuppert, Christian Karagiannidis

## Abstract

**Background:** Data on long-term outcomes of hospitalized COVID-19 patients are scarce.

**Objective:** To provide a detailed account of hospitalized COVID-19 patients until 180 days after their initial hospitalization.

**Design:** Nationwide cohort study using claims data from the German Local Health Care Funds, the health insurer of one-third of the German population.

**Setting:** Germany.

**Patients:** Adult patients hospitalized in Germany between Feb 1 and April 30, 2020 with PCR-confirmed COVID-19 and a related principal diagnosis.

**Measurements:** Patient characteristics and ventilation status, in-hospital, 30-, 90- and 180- day mortality measured from admission, and 180-day readmission measured from discharge. Multivariable logistic regression model of independent risk factors for 180-day mortality.

**Results:** Of 8679 patients (median age, 72 years), 2161 (24.9%) died during the index hospitalization. 30-day mortality was 23.9% (2073/8679), 90-day mortality 27.9% (2425/8679), and 180-day mortality 29.6% (2566/8679). The latter was 52.3% (1472/2817) for patients aged ≥ 80 years, and 53.0% for patients who had been ventilated invasively (853/1608). Risk factors for 180-day mortality included coagulopathy, BMI ≥ 40, and age. Female sex was a protective factor. Of 6235 patients discharged alive, 1668 patients were readmitted a total of 2551 times within 180 days, resulting in an overall readmission rate of 26.8%.

**Limitations:** We could not stratify patients by ICU treatment as it is not coded separately. Furthermore, we cannot exclude residual confounding in our analysis of risk factors nor determine causality between risk factors and long-term mortality given the observational nature of this study.

**Conclusion:** This nationwide cohort study of hospitalized COVID-19 patients in Germany found considerable long-term mortality and readmission rates, especially among patients with coagulopathy. Close follow-up after hospital discharge may improve long-term outcome.

## Introduction

Within one year, the SARS-CoV-2 pandemic has affected more than 125 million people worldwide. During the first wave in spring 2020, hospitalization rates were high, reaching up to 70% in France, 55% in Spain, 50% in the UK and 20% in Germany until the end of April (1-4). Mortality rates of hospitalized patients were also high at more than 20% in France, United Kingdom and Germany, especially in patients requiring mechanical ventilation with up to 50% (1-5). However, little is known about the long-term outcome of those COVID-19 patients who were discharged alive, particularly in regard of mortality, hospital readmissions and long-term complications. Furthermore, there is increasing evidence of a long-COVID syndrome, affecting different organ systems, lasting from primary neurological symptoms to non-resolving structural lung diseases.

Recently, the 6-month follow-up data of the first 1733 hospitalized COVID-19 patients from Wuhan were published (6). COVID-19 survivors suffered mainly from fatigue or muscle weakness, sleep difficulties, and anxiety or depression. Furthermore, patients who were more severely ill showed reduced pulmonary diffusion capacities and abnormal chest imaging. These data are in line with recent data of an increased risk for neurological and psychiatric disorders after 6 months (7-11).

In regard of severely ill COVID-19 patients, it has been shown in 167 COVID-19 patients admitted to an intensive care unit (ICU) in Wuhan that older age and thrombocytopenia among others were shown to be a significant risk factors for mortality at 28, 90 and 180 days (12). Furthermore, in a multicenter-study from five European countries including 132 hospitalized COVID-19 patients requiring extracorporeal membrane oxygenation (ECMO), also advanced age and low pH before ECMO were associated with increased risk of six-month mortality, which was overall 53% (70/132) (13). To date, no study has evaluated 6-month readmission.

For large cohorts outside Wuhan, follow-up data have been reported up to a maximum of three months. 90-day *post-admission* outcomes have been reported from various European countries, albeit often for single hospitals. 90-day mortality ranged from 11% in Spain (14) to 29% in Denmark (15) (both single-center studies) for all hospitalized patients, and was 27% in Sweden (16), 31% in Belgium, France and Switzerland (both multi-center studies) (17) and 35% in a Danish single-center study for patients treated on the ICU (15). An analysis of the United States’ Centers for Disease Control and Prevention of more than 100 000 hospitalized COVID-19 patients for a 6-month period *post-discharge* revealed a readmission rate of 9% to the same hospital (18).

Since large cohort 6-month follow up data are currently lacking, the aim of this first nationwide cohort study was to determine detailed characteristics and outcomes of hospitalized COVID-19 patients with completed hospital treatments in a large, unselected and unbiased cohort of patients with confirmed COVID-19 diagnosis in one of the least resource limited health care systems, particularly focusing on patients requiring mechanical ventilation. Outcomes of interest were mortality within 180 days of initial admission and readmission to any hospital within 180 days of discharge. Patient-related risk factors for 180-day mortality were investigated.

## Methods

### Data Sources

We use anonymized nationwide administrative claims data from the German local health care funds (Allgemeine Ortskrankenkassen, AOK), the largest sickness fund group within Germany’s statutory health insurance system. AOK provides statutory health insurance for roughly 32 percent of the German population (19). Membership is open to anyone regardless of factors such as professional affiliation, income, age or comorbidities (20). Of note, nearly every inhabitant of Germany has an obligatory health care insurance. According to the German accounting method for the health care system, all diagnoses, outcomes and procedures must be reported to the sickness funds, as required by law. Hence, the data set includes detailed information on patient characteristics such as age, gender, length of hospital stay, diagnoses and procedure codes and discharge type (survival status). Diagnoses were coded according to the 10^th^ revision of the International Classification of Diseases (ICD-10-GM) and procedures according to the International Classification of Procedures in Medicine, the “Operationen- und Prozedurenschlüssel” (OPS). Further core data of the health care funds include insurance status and overall survival status.

For the analyses, we included only patients with a confirmed COVID-19 infection by reverse transcription polymerase chain reaction (PCR) (diagnosis code U07.1) who are at least 18 years old and were admitted to hospital between February 1, 2020 and April 30, 2020 inclusive, and were discharged by June 30, 2020. As COVID-19 cannot be coded as a principal diagnosis, which is defined as the main reason for hospitalization, we cannot distinguish between patients who have been hospitalized for COVID-19 or with COVID-19 for another reason. Therefore, we limited the analysis to those patients with a COVID-19-related principal diagnosis of respiratory failure, pulmonary embolism, viral infection, sepsis or renal failure during this initial “index” hospitalization. Furthermore, because one insured person might have had several hospital stays during the observation period due to a transfer from one hospital to another, we grouped patients with adjacent completed hospital stays into one case.

### Variables

Death was measured as in-hospital, i.e. occurring during the index hospitalization, as well as 30 days, 90 days and 180 days after the day of initial admission. As we were interested in 30-, 90- and 180-day mortality post initial admission date, patients had to be continuously insured with AOK for that period of time, unless they died earlier while still insured with AOK.

For readmissions, we followed patients discharged alive from the index hospitalization for 180 days after the day of discharge. The discharge date was chosen for the patients’ length of index hospital stay varied widely. We determined both how many patients were readmitted at least once, and how many readmissions occurred in total for any cause and for readmission with potentially COVID-19 related systemic, respiratory, renal and neurological, gastrointestinal and liver as well as cardiovascular diagnoses only.

### Statistical Analysis

We analyzed and report outcomes for patients grouped according to their characteristics during the index hospitalization, namely, gender, age groups (18–59 years, 60–69 years, 70– 79 year, ≥80 years), selected Elixhauser comorbidities (21, 22), and whether or not they received any form of mechanical ventilation. For ventilated patients, two subgroups were formed: (a) patients with non-invasive mechanical ventilation only (NIV), and (b) patients with invasive mechanical ventilation (IMV), including those with non-invasive mechanical ventilation failure. In addition, we report on other procedures, such as dialysis or ECMO, as well as complications, such as septic shock, acute respiratory distress syndrome (ARDS), or renal failure post medical procedures. For continuous variables, we report means and standard deviation (SD), and medians and inter-quartile ranges (IQR). For categorical variables, we report percentages. All variables are reported for patients with and without mechanical ventilation and for the different ventilation groups. Kaplan-Meier curves for survival up to 180 days after the initial admission are presented by gender, age, type of ventilation, and selected Elixhauser comorbidities. Multivariable logistic regression with cluster-robust standard errors was used to model the odds of 180 day-mortality after admission as function of age, sex, body mass index (BMI) categories (30≤34, 35≤39, ≥40 kg/m^2^) and Elixhauser comorbidities. A model including all potential risk factors was estimated first, with subsequent removal of risk factors which did not prove statistically significant (p ≥ 0.05). A Wald test was performed to confirm that removing these risk factors from the respective model did not result in a substantial loss in model fit. Adjusted odds ratios (OR) and 95% confidence intervals (CIs) were calculated. Due to non-parallel Kaplan-Meier curves, no Cox-proportional hazard model could be estimated. All analyses were performed using STATA 16.0 (StataCorp, College Station, Texas).

The study was approved by the Ethics Committee of the Witten/Herdecke University (research ethics board number 92/2020).

## Results

We identified 11 459 patients with PCR-confirmed COVID-19 diagnosis who were first admitted as inpatients from February 1 to April 30, 2020 and discharged before June 30, 2020. We excluded 2780 patients who did not suffer from a COVID-19-related principal diagnosis (n = 2572) or who could not be followed for the full six month-period after their initial admission or until death (n = 208). In total, 8679 patients were confirmed eligible and included in the analysis.

### Demographics and Comorbidity

The patients’ demographic characteristics are depicted in Table 1. The cohort comprised of slightly more men (4641/8679; 53.5%) than women (4038/8679; 46.5%). Median age was 72 years (IQR 57 to 82), the largest share of patients being in the age group of 80 years and older (2817/8679; 32.5%). The most frequent comorbidities were hypertension (4920/8679; 56.7%), fluid and electrolyte disorders (4641/8679; 53.5%), diabetes mellitus (uncomplicated: 1912/8679; 22.0%; complicated: 734/8679; 8.5%), cardiac arrhythmia (2373/8679; 27.3%), renal failure (1994/8679; 23.0%), and congestive heart failure (1652/8679; 19.0%).

**Table 1.**
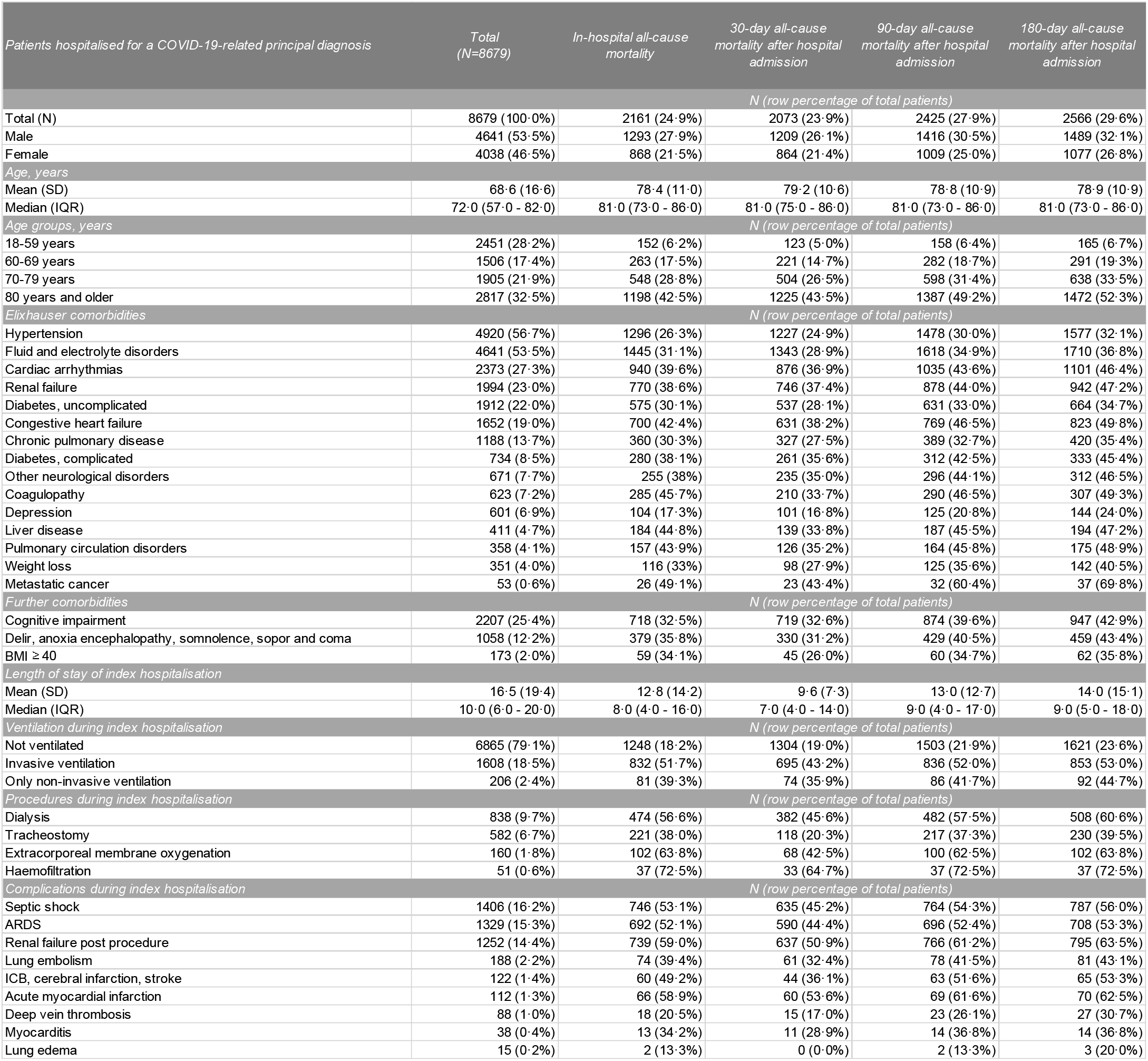
Patient characteristics. Data are n (%), median (IQR) or mean (SD). BMI: Body Mass Index; ARDS: acute respiratory distress syndrome

### Index Hospitalization

Median length of stay of the index hospitalization was 10 days (IQR 6 to 20); mean length of stay was 16.5 (SD 19.4) days (Table 1). 6865 (79.1%) patients were treated without mechanical ventilation and 1814 (20.9%) were treated with mechanical ventilation, of whom 1608 (18.5%) received IMV and 206 (2.4%) received NIV only. The patients’ demographic characteristics stratified by ventilation status can be found in Appendix Table 1. Dialysis was performed in 9.7% (838/8679) and ECMO in 1.8% (160/8679). Frequent complications during the index hospitalization included septic shock (1406/8679; 16.2%), ARDS (1329/8679; 15.3%), and acute renal failure (1252/8679; 14.4%).

### All-cause Mortality

Of the 8679 included patients, 2161 (24.9%) died during the index hospitalization (Table 1). Measured from day of initial admission, 30-day all-cause mortality was 23.9% (2073/8679), 90-day all-cause mortality was 27.9% (2425/8679), and 180-day all-cause mortality was 29.6% (2566/8679).

Women’s survival rates were about 5%-points higher than men’s at all three time points (Figure 1a). Survival was also strongly associated with age. Patients in the age group of 18-59 years had a 180-day mortality of 6.7% (165/2451), which increased to 19.3% (291/1506) in patients aged 60-69 years, 33.5% (638/1905) in patients aged 70-79, and 52.3% (1472/2817) in patients aged 80 years or above (Figure 1b). The relative difference between in-hospital and 180-day all-cause mortality was also greatest in this age group with an additional 9.7% of patients who died during follow-up (Table 1). Patients who were not ventilated had better survival than ventilated patients (Figure 1c), their 180-day mortality being 23.6% (1621/6865) compared to 52.1% (945/1814). Amongst ventilated patients 30-, 90- and 180-day mortality rates were about 8%-points lower in patients treated with NIV compared to IMV.

**Figure 1.**
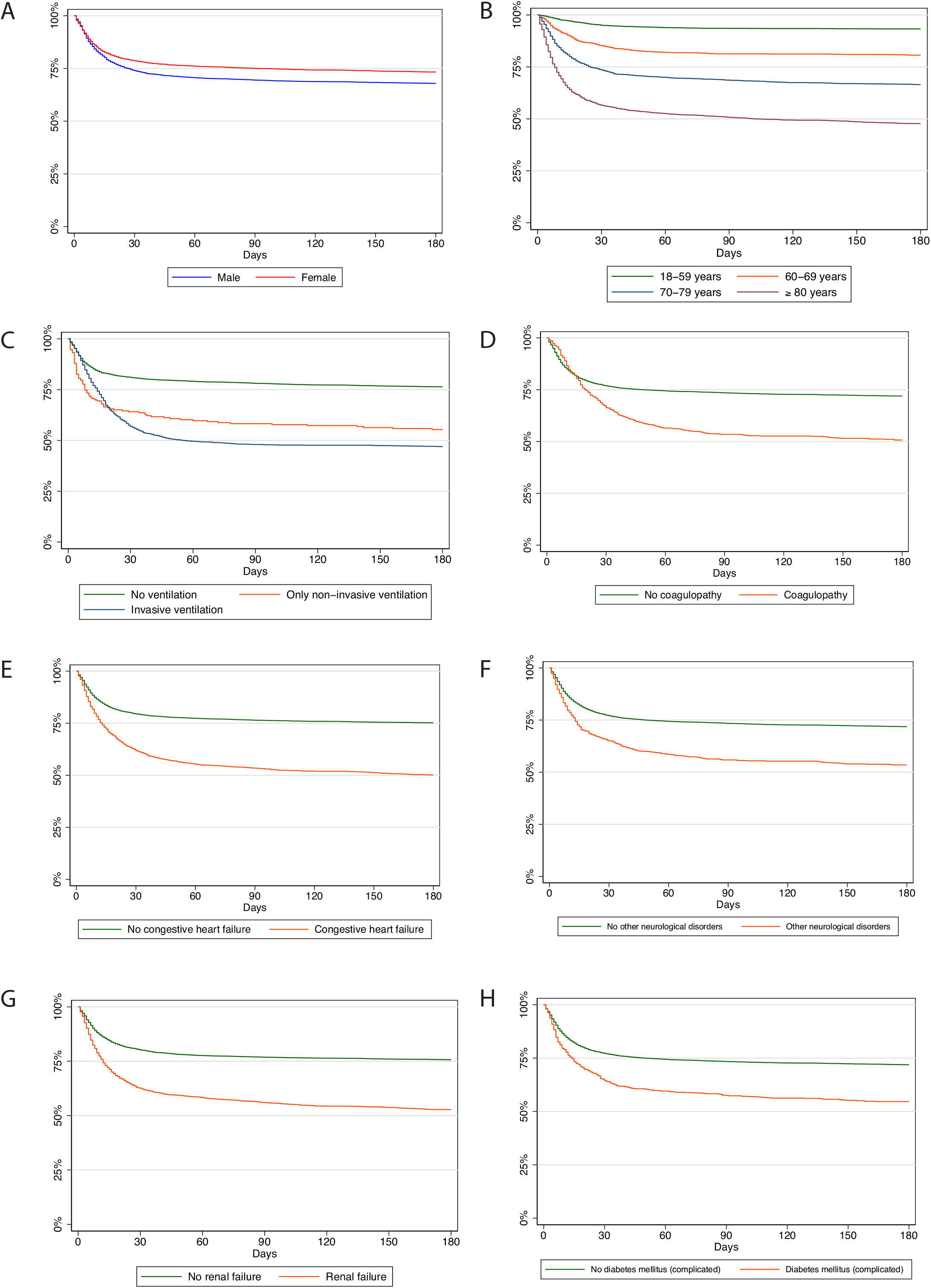
Kaplan-Meier survival curves of all hospitalized COVID-19 patients followed for 180 days after hospital admission.

Among the comorbidities present in at least 5% of the sample, the greatest difference between 30-day and 180-day mortality was observed for the following comorbidities: Coagulopathy, congestive heart failure, other neurological disorders, renal failure, and complicated diabetes mellitus (Table 1). Patients with coagulopathy had the largest increase in 180-day mortality, with an additional 15.6% (97/623) of patients who died between 30 and 180 days (Figure 1d). Survival was considerably better for patients not having these respective comorbidities. For patients with coagulopathy, 180-day mortality was 49.3% (307/623; Figure 1d), with congestive heart failure 49.8% (823/1652; Figure 1e), with other neurological disorders 46.5% (312/671; Figure 1f), with renal failure 47.2% (942/1994; Figure 1g), and with complicated diabetes 45.4% (333/734; Figure 1h).

In a subgroup-analysis of ventilated patients only, the observed trends regarding gender, age, and ventilation type (non-invasive/invasive) persisted (Figure 2a-c), while the differences narrowed (congestive heart failure and renal failure; Figure 2e, g) or vanished in the analyses per comorbidity (Figure 2d, f, h). For patients with coagulopathy, the 30-day mortality was even lower than that of patients with no coagulopathy (Figure 2d); for patients with other neurological disorders, this was even observed beyond day 90 (Figure 2f).

**Figure 2.**
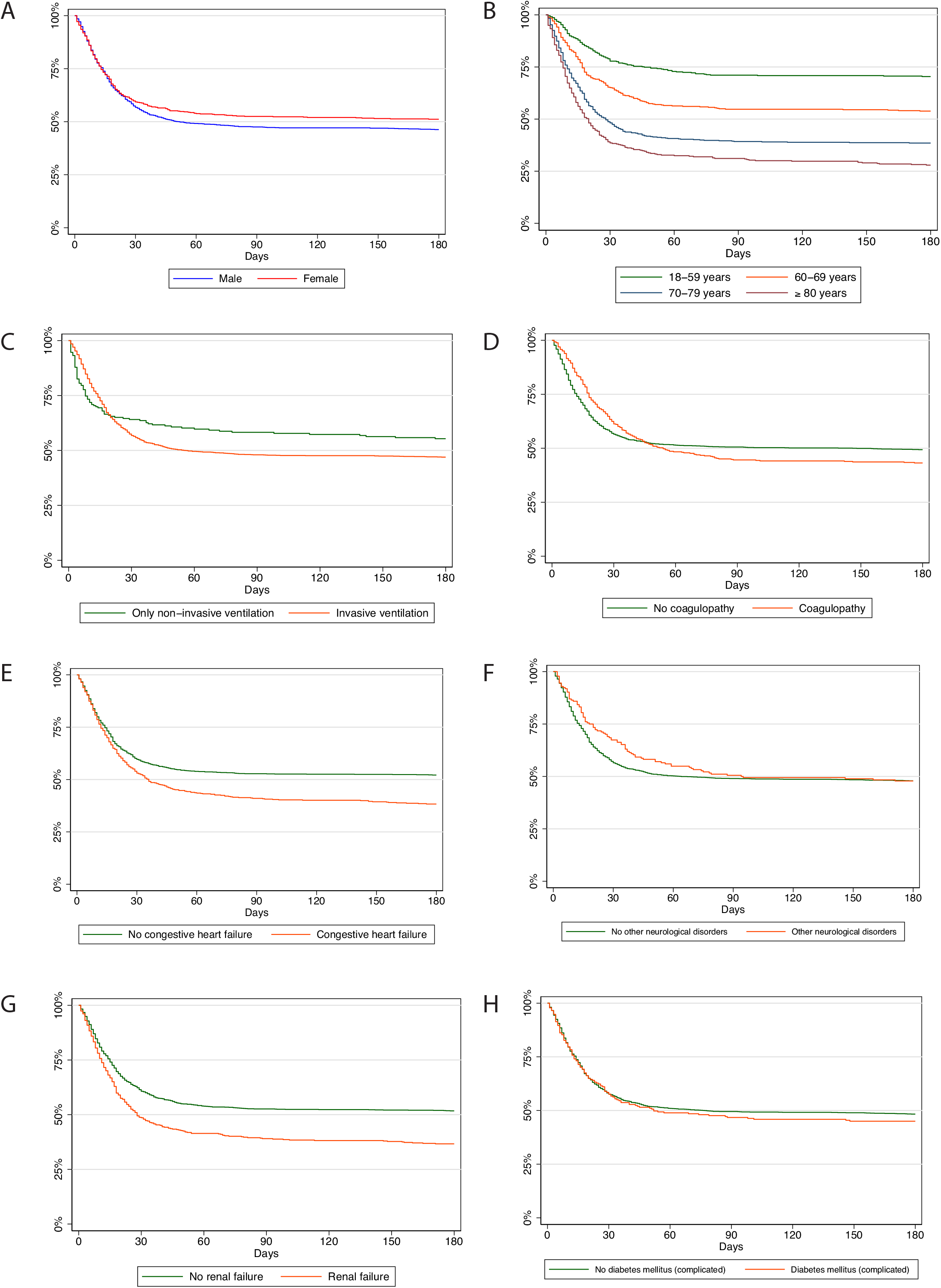
Kaplan-Meier survival curves of all hospitalized COVID-19 patients on mechanical ventilation followed for 180 days after hospital admission.

The results of the logistic regression model are presented for all covariates significantly associated with 180-day mortality in Figure 3. Adjusted for age, sex and comorbidities, strong associations with increased odds of 180-day mortality (OR > 2) were observed for patients with a BMI ≥ 40 (OR 2.01, 95%-CI 1.33 to 3.05), liver disease (OR 2.45, 95%-CI 1.85 to 3.25), metastatic cancer (OR 8.02, 95%-CI 3.57 to 18.00), and coagulopathy (OR 2.31. 95%-CI 1.82 to 2.94). For age, the OR was 1.08 per year, indicating that the odds for 180-day all-cause mortality increase by the factor 2.21 per additional 10 years of age (1.082449^10), 4.88 per additional 20 years of age (1.082449^20), and so on. Conversely, a strong association for decreased odds of 180-day mortality was observed for female patients (OR 0.63, 95%-CI 0.56 to 0.70), and patients with depression (OR 0.46, 95%-CI 0.37 to 0.57).

**Figure 3.**
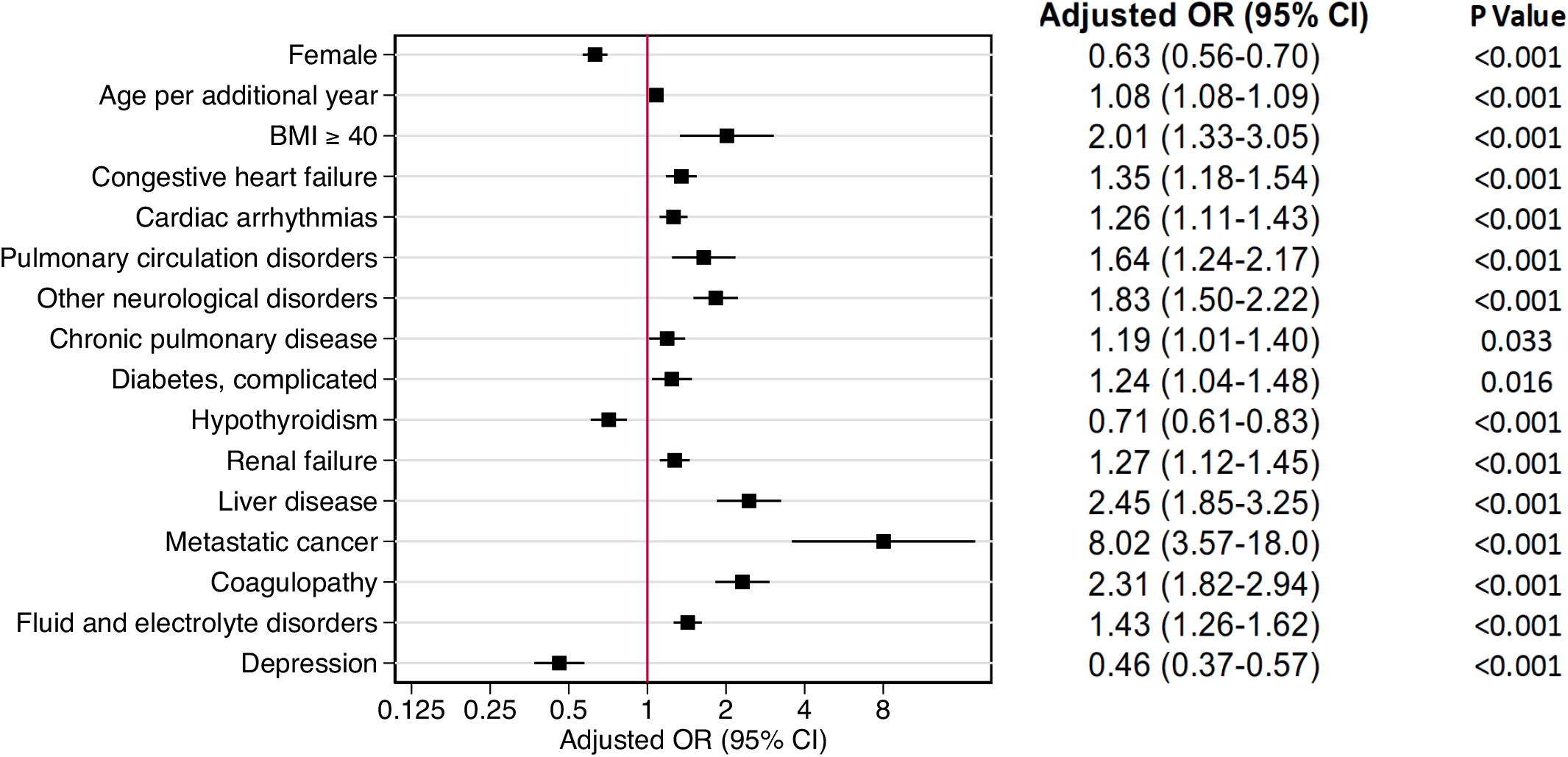
Multivariable logistic regression analysis for 180-days all-cause mortality after hospital admission. CI confidence interval, OR odds ratio, BMI body mass index. Only significant risk factors are included in model.

### Readmission

Of all 6518 patients discharged alive from the index hospitalization, 283 patients were lost to follow-up for they died after 180 days of initial admission but before 180 days post discharge, or were not insured with AOK anymore. Of the remaining 6235 patients, 1668 were readmitted a total of 2551 times for any cause within 180 days post discharge, resulting in an overall readmission rate of 26.8%.

405 (6.2%) patients of the 6518 patients discharged alive died within 180 days from initial admission. Around half of those (201/405; 49.6%) were readmitted within 180 days from discharge, and the other half not (204/405; 50.4%). Thus, the increase in 180-day mortality post discharge from the initial hospitalization was much higher in readmitted patients (201/1668; 12.1%) than in those not readmitted (204/4850; 4.2%).

Unlike mortality rates, readmission rates were only slightly higher amongst men (893/3204; 27.9%) than women (775/3031; 25.6%) (Table 2). Patients treated with IMV (231/717; 32.2%) had higher readmission rates than patients treated with NIV (34/121; 28.1%) or patients who were not ventilated (1403/5397; 26.0%).

**Table 2.** Readmission within 180-days of discharge.

| *Readmission among patients discharged alive from index hospitalisation\** | *Total*<br>*(N=62 35)* | *Male*<br>*(N=3204)* | *Female*<br>*(N = 3031)* |
| --- | --- | --- | --- |
| All patients with at least one readmission | 1668 (26.8%) | 893 (27.9%) | 775 (25.6%) |
| Patients with ventilation during index admission |  |  |  |
| Exclusively non-invasive ventilation | 34/121 (28.1%) | 20/78 (25.6%) | 14/43 (32.6%) |
| Invasive ventilation | 231/717 (32.2%) | 154/453 (34.0%) | 77/264 (29.2%) |
| No ventilation | 1403/5397 (26.0%) | 719/2673 (26.9%) | 684/2724 (25.1%) |
| *COVID-19-related readmission within 180 days* |  |  |  |
|  | *Principal or secondary diagnosis*<br>*% of readmitted patients* |  |  |
| Any systemic/ respiratory/ renal/ neuro/ gastrointestinal and liver/ cardiocascular/ ventilation disorder or complication/ COVID-19 | 1011 (60.6%) | 550 (61.6%) | 461 (59.5%) |
| COVID-19 (U07.1!) | 212 (12.7%) | 124 (13.9%) | 88 (11.4%) |
| Systemic disorders/complications | 162 (9.7%) | 104 (11.6%) | 58 (7.5%) |
| Respiratory disorders/complications | 601 (36.0%) | 351 (39.3%) | 250 (32.3%) |
| Renal disorders/complications | 203 (12.2%) | 119 (13.3%) | 84 (10.8%) |
| Neurological disorders/complications | 490 (29.4%) | 244 (27.3%) | 246 (31.7%) |
| Gastrointestinal and liver disorders/complications | 117 (7.0%) | 77 (8.6%) | 40 (5.2%) |
| Cardiovascular disorders/complications | 192 (11.5%) | 115 (12.9%) | 77 (9.9%) |
| Mechanical ventilation (non-invasive and invasive) | 103 (6.2%) | 66 (7.4%) | 37 (4.8%) |
| * patients followed until 180 days post discharge only |  |  |  |

The majority of the readmitted patients (1011/1668; 60.6%) had potentially COVID-19-related systemic, respiratory, renal, neurological, gastrointestinal and liver, cardiovascular principal/secondary diagnoses or were ventilated at readmission (Table 2). When viewed separately, the most frequent principal or secondary diagnoses for readmission were respiratory disorders or complications (601/1668; 36.0%), and neurological disorders or complications (490/1668; 29.4%). Less than ten percent of the patients received mechanical ventilation at readmission (103/1668; 6.2%). 212 (12.7%) patients tested again positive for COVID-19 at readmission.

## Discussion

This is the first nationwide study showing the 6-month outcome of hospitalized COVID-19 patients in an unselected cohort, including readmissions within this time period. The major findings are the high 6-month mortality in COVID-19 patients in particular in those requiring mechanical ventilation with 53%, that women have a sustained and profound beneficial 6-month outcome compared to men, and that patients in the age above 80 years have the worst outcome with a mortality rate of 52% or up to 71% for those being ventilated. Furthermore, a poor long-term outcome is associated with coagulopathy, liver diseases or severe obesity. Last, readmission rates reached 27%, of whom most of the patients with potentially COVID-19 related diagnoses were admitted for respiratory or neurological disorders or complications.

The initial in-hospital mortality of 25%, which is in line with many other European countries, increased to an all-cause mortality of 30% after 6 months, which demonstrates severe major prolonged implications of this disease, rather more than we would have expected. It is noticeable that ventilated patients have a poor overall outcome, especially patients over 70 years of age. In contrast it is also evident that in the younger age groups there is only a slight increase in mortality after hospital discharge, although serious long-term consequences, having a huge impact on morbidity and quality of life may occur. In regard of risk factors being associated with a poor or a more beneficial long-term outcome several key factors became obvious. Overall, women show a better long-term outcome than men regardless of other confounding factors in terms of mortality, which may be due to the more beneficial immune response compared to men (23). On the other hand, factors contributing to increased mortality are especially disorders of the coagulation system, or liver disease, as already known for in-hospital mortality. These data are in line with our current understanding of COVID, in particular disorders of the coagulation or cerebrovascular system (24-26).

While diabetes generally worsens the outcome, this is not the case for patients being mechanically ventilated, whereas acute renal failure, congestive heart failure and age account for a worse outcome independent of being ventilated. However, further data on patient’s diagnosis before admission with COVID-19 are missing in this study. In the light of the current analysis, it should be critically evaluated whether current intensive care therapy, including mechanical ventilation in patients over 80 years of age, is really effective in view of the very high mortality rate, or whether, in the future, we should develop narrow criteria for eligible patients who could have a more favorable outcome. This certainly includes the frailty of the elderly (27-29).

Within 180 days of discharge, there was a high number of readmissions with 27% of those discharged alive independent of gender, primarily due to respiratory or neurological diagnoses. Also, mortality in readmitted patients remains rather high with 6% of all discharged patients. Beside the lung, readmissions with neurological complications maybe a severe manifestation of the disease which may prolong for several months and lead to long-term complications, actually unknown if resolving or not. Of not, patients with coagulopathy had the highest 180-day mortality, which sheds a special light on early detection of possible thrombosis or pulmonary embolism following the initial admission. In general, close follow-up by general practitioners and the corresponding specialized disciplines is needed for early interventions regarding neurocognitive impairments. Of note, 13% of all readmitted patients were still or again positive for SARS-CoV-2 pointing out, that virus elimination may take a long time in some severely ill patients.

One limitation of our study is the data source, which only includes patients from the one group of German sickness funds. However, it is the largest group which accounts for 1/3 of the total population, providing a large sample representative for the German population, even if the very old are somewhat overrepresented (20). Hospital data is of high quality because disease codes (ICD) and procedure codes (OPS) are relevant for the amount of remuneration and therefore verified by hospitals and sickness funds. Second, patient-specific data are limited to inpatient diagnoses, procedures and initial characteristics, so some pre-existing conditions might be unknown. Third, we stratified by mechanical ventilation, but not by ICU treatment (as it is not coded separately), which sometimes includes high-flow oxygen therapy without mechanical ventilation. Fourth, detailed data such as laboratory values and information on patient preferences or clinical decision making that may impact the initiation of invasive treatments are not available. Fifth, given its observational nature, this study cannot determine causality between risk factors and long-term mortality.

In this nationwide cohort of patients hospitalized for COVID-19 considerable long-term mortality and readmission rates were observed. Patients with coagulopathy had the largest increase in 180-day mortality, followed by congestive heart failure, neurological diseases and acute renal failure. However, female sex is the profound protective factor in COVID-19 disease.

## Data Availability

Data access upon personal discussion.

## Author contributions

CG, RB, and CK conceived the study and its design. CG and MS had full access to the data, have verified all data, and take responsibility for the integrity of the data and accuracy of the analysis. CG, RB, MS, TR, GS and CK contributed to data analyses. CG, RB, GS, WH, SW-C, AS, and CK contributed to data interpretation. RB, TR and CK wrote the main draft of the manuscript. All authors revised the main draft critically for important intellectual content and contributed equally to the final draft of the manuscript. All authors have read and approved the submitted manuscript and take full responsibility for the integrity of this work.

