## Supplemental Figure 1 for "6-Month Follow Up of 8679 Hospitalized COVID-19 Patients in Germany: A Nationwide Cohort Study"

Supplemental Table 1: Patient characteristics. Data are n (%), median (IQR) or mean (SD). BMI: Body Mass Index; ARDS: acute respiratory distress syndrome

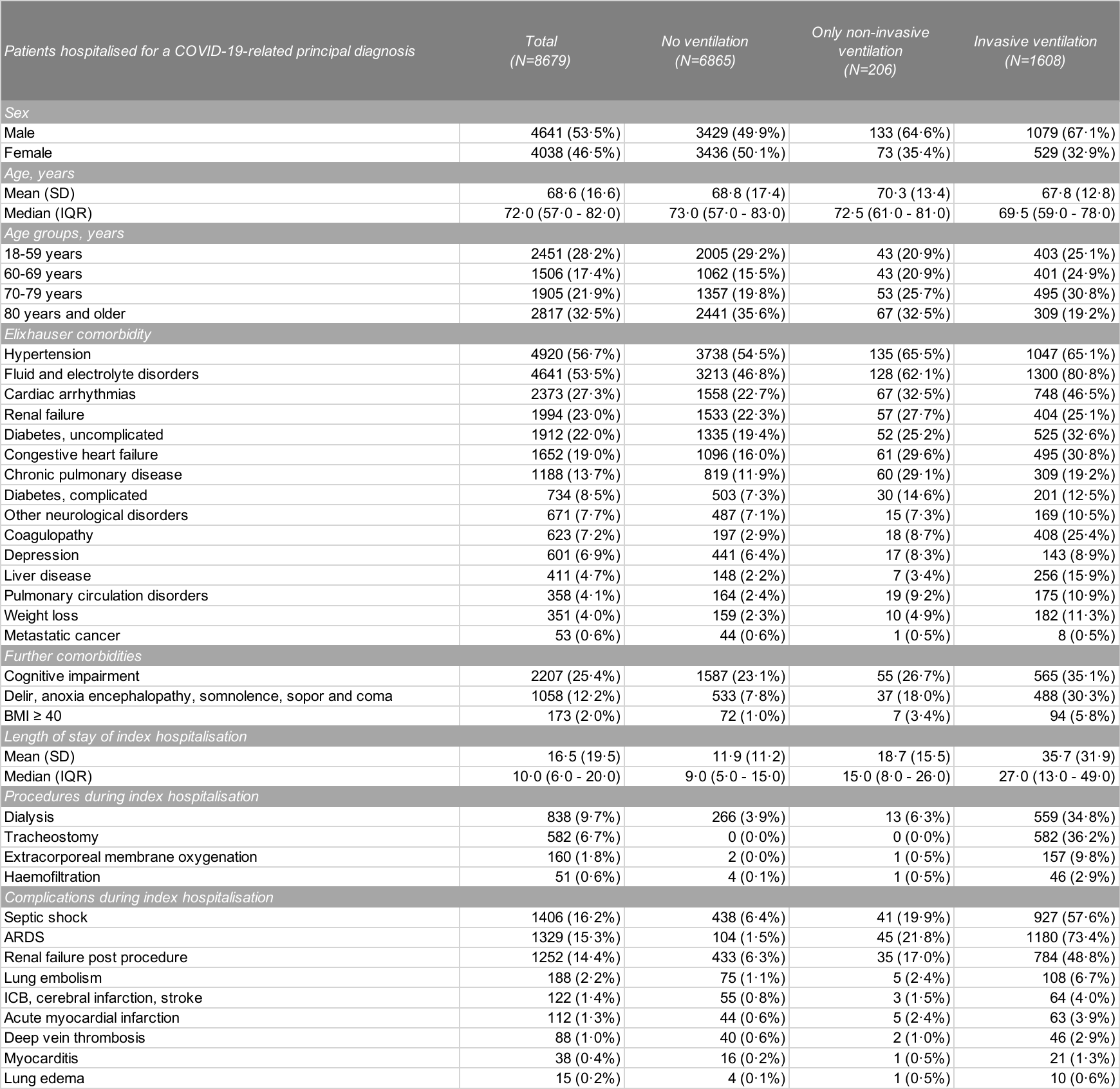
